# Cortical thickness and sub-cortical volumes in post-H1N1 narcolepsy type 1: A brain-wide MRI case-control study

**DOI:** 10.1101/2023.03.13.23287231

**Authors:** Hilde T. Juvodden, Dag Alnæs, Ingrid Agartz, Ole A. Andreassen, Andres Server, Per M. Thorsby, Lars T. Westlye, Stine Knudsen-Heier

## Abstract

**Objective:** There was an increased incidence of Narcolepsy type 1(NT1) after the H1N1-mass vaccination in 2009/2010 which has been associated with the Pandemrix®-vaccine. We performed the first case-control comparison of MRI-based global and sub-cortical volume and cortical thickness in post-H1N1(largely Pandemrix®-vaccinated) NT1 patients compared with healthy controls.

**Methods:** We included 54 post-H1N1 NT1 patients (51 with confirmed hypocretin-deficiency; 48 H1N1-vaccinated with Pandemrix®; 39 females, mean age 21.8 ± 11.0 years) and 114 healthy controls (77 females, mean age 23.2 ± 9.0 years). 3T MRI brain scans were obtained, and the T1-weighted MRI data were processed using FreeSurfer. Group differences among three global and 10 sub-cortical volume measures and 34 cortical thickness measures for bilateral brain regions were tested using general linear models with permutation testing. We corrected for multiple testing with the Benjamini-Hochberg procedure with the false discovery rate at 5%.

**Results:** Patients had significantly thinner brain cortex bilaterally in the temporal poles (*Cohen’s d*=0.68, *p*=0.00080), entorhinal cortex (*d*=0.60, *p*=0.0018) and superior temporal gyrus (*d*=0.60, *p*=0.0020) compared to healthy controls. The analysis revealed no significant group differences for sub-cortical volumes.

**Conclusions:** Post-H1N1(largely Pandemrix®-vaccinated) NT1 patients have significantly thinner cortex in temporal brain regions compared to controls. We speculate that this effect can be partly attributed to the hypothalamic neuronal change in NT1, including loss of function of the widely projecting hypocretin-producing neurons and secondary effects of the abnormal sleep-wake pattern in NT1. Alternatively, the findings could be specific for post-H1N1 (largely Pandemrix®-vaccinated) NT1 patients.

## 1. Introduction

Narcolepsy type 1 (NT1) is a neurological sleep disorder characterized by cataplexy, excessive daytime sleepiness and disturbed night sleep, and it is also common for the patients to experience hypnagogic hallucinations and sleep paralysis. [1] NT1 patients have a loss/dysfunction of hypocretin-producing neurons in the hypothalamus most likely due to an autoimmune process. [1–4] Hypocretin-deficiency can be measured in the cerebrospinal fluid and used diagnostically. [1] The hypocretin-producing neurons are central in sleep/wake-regulation, they project widely to other brain regions, and the loss of these neurons have been shown to disrupt the functioning of multiple frontal, limbic, diencephalic and brainstem networks. [1, 5]

The incidence of narcolepsy increased by 10-fold after the H1N1-mass vaccination in 2009/2010 in Norway and several other countries. [6, 7] The risk associated with H1N1-vaccination has in several studies been shown to be limited to the AS03-adjuvated Pandemrix® vaccine. [7, 8] Pandemrix was administered in large-scale with over 31 million doses to populations in Europe, including Norway, Finland, Germany, Ireland, France, United Kingdom and Sweden. [7, 8] In China an increase in narcolepsy cases was detected after the H1N1-infection in the absence of vaccination [9], but a similar increase without vaccination have not been found in other countries. [1]

A recent MRI T1-study by our group [10] on the same 54 post-H1N1 patients and 114 controls, have explored group differences in volume in a segmentation of the hypothalamus and we therefore wanted to explore the possible distributed nature of structural affection in the rest of the brain of NT1 patients, to gain insight into the disease mechanisms and possible consequences due to an abnormal sleep-wake pattern. Previous structural brain-wide MRI studies investigating volume in narcolepsy patients compared to controls have reported conflicting results, [11–22] possibly due to relatively small and heterogenous patient samples.

The current study is the first study to obtain a brain-wide assessment of cortical thickness and subcortical volumes in a post-H1N1, largely Pandemrix® -vaccinated NT1 patient sample as well as the largest brain-wide T1-study of NT1 patients to date. Due to the conflicting results from previous studies and our study representing the first study of a largely Pandemrix®-vaccinated NT1 patient sample, we wanted to perform a brain-wide screening for regions of interest, obtaining three global and 10 bilateral sub-cortical volume measures and 34 bilateral cortical thickness measures covering the full brain.

## 2. Materials and Methods

### 2.1 Participants

We recruited 70 suspected NT1 patients from June 2015 to April 2017 with disease onset after the 2009/2010 H1N1-pandemic and vaccination campaign. The patients were a part of a national post-H1N1 narcolepsy follow-up project initiated by the Norwegian Ministry of Health and Care Services. Patient and their families were referred for narcolepsy disease education and counseling courses at the Norwegian Centre of Expertise for Neurodevelopmental Disorders and Hypersomnias (NevSom) and were invited to also participate in the scientific part of the project during their stay at our center. Before inclusion written informed consent was provided by all participants (parents signed for children <16 years of age) and the study was approved by the Norwegian regional committees for medical and health research ethics (REK).

Exclusion criteria for patients were severe somatic, neurological or psychiatric disorders, metallic implants, neuroradiological findings requiring a clinical follow-up and previous head injury with loss of consciousness for 10 minutes or 30 minutes amnesia.

Two patients were excluded due to severe psychiatric/neurological disorders, two patients were excluded due to not fulfilling the NT1 ICSD-3 diagnostic criteria, [23] four patients were excluded due to dental braces leading to bad quality MRI data, and eight patients due to excessive head movement during MRI. The remaining 54 patients were included in the analysis. The clinical and demographic information are summarized in Table 1. The average Apnea Hypopnea Index (AHI) for the patients were 3.3 ± 9.1. and six patients had AHI ≥ 5. Patients were requested to pause all narcolepsy medication and other medication affecting sleep/wake for 14 days prior to inclusion, however three patients were unable to do this completely. One patient only paused Venlafaxine for 7 days prior to inclusion due to severe cataplexy; and due to comorbidity two other patients continued to use Fluoxetine and Catapresan, respectively.

**Table 1.**
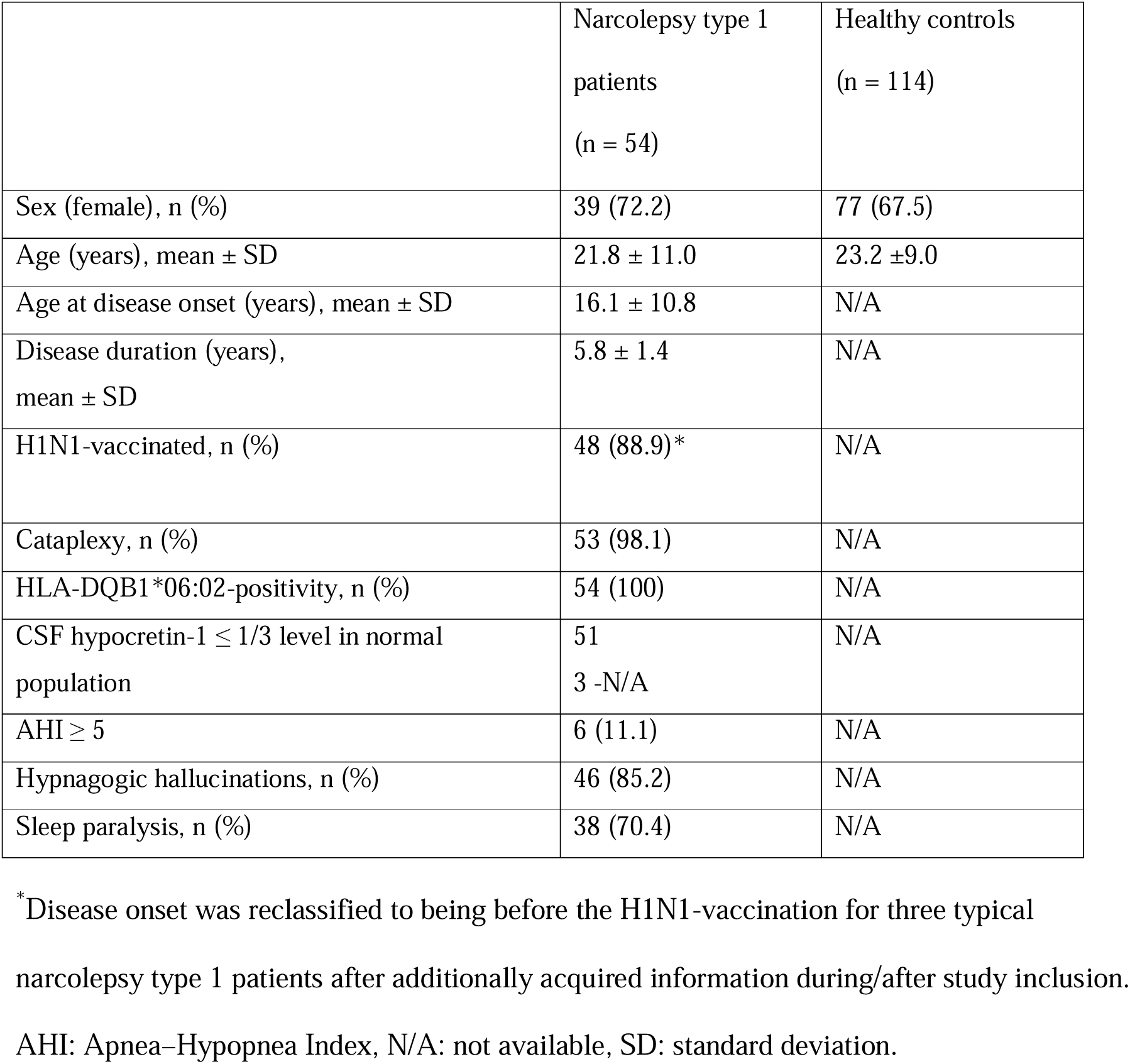
Demographic and clinical data.

|  | Narcolepsy type 1<br>patients<br>(n = 54) | Healthy controls<br>(n = 114) |
| --- | --- | --- |
| Sex (female), n (%) | 39 (72.2) | 77 (67.5) |
| Age (years), mean ± SD | 21.8 ± 11.0 | 23.2 ± 9.0 |
| Age at disease onset (years), mean ± SD | 16.1 ± 10.8 | N/A |
| Disease duration (years),<br>mean ± SD | 5.8 ± 1.4 | N/A |
| H1N1-vaccinated, n (%) | 48 (88.9)* | N/A |
| Cataplexy, n (%) | 53 (98.1) | N/A |
| HLA-DQB1*06:02-positivity, n (%) | 54 (100) | N/A |
| CSF hypocretin-1 ≤ 1/3 level in normal<br>population | 51<br>3 -N/A | N/A |
| AHI ≥ 5 | 6 (11.1) | N/A |
| Hypnagogic hallucinations, n (%) | 46 (85.2) | N/A |
| Sleep paralysis, n (%) | 38 (70.4) | N/A |

\*Disease onset was reclassified to being before the H1N1-vaccination for three typical narcolepsy type 1 patients after additionally acquired information during/after study inclusion.
AHI: Apnea–Hypopnea Index, N/A: not available, SD: standard deviation.

Additional information obtained during or after study inclusion revealed the disease onset to be prior to the H1N1-vaccination campaign for three patients in the H1N1-vaccinated group. These three patients had typical NT1 phenotypes with cataplexy, HLA-DQB1*06:02-positivity and hypocretin-deficiency and hence remained included in the study. The H1N1-vaccination information was obtained from the official Norwegian Immunisation Registry (SYSVAK). The only H1N1-vaccine used in Norway was Pandemrix®. Two patients in the H1N1-vaccinated group were not registered in SYSVAK but reported having been H1N1-vaccinated at their workplace.

Comorbidities are common in NT1 patients [2] and we therefore included the following comorbidities in the main analysis: migraine (n=6), Asperger syndrome (n=2), attention-deficit hyperactivity disorder (ADHD; n=1), anxiety (n=1), depression (n=1), Tourette syndrome (n=1), kidney disease (n=1), type 2 diabetes (n=1), hypothyroidism (n=1) and premature birth without severe long-term complications (n=1). There were in total 14 patients with these comorbidities as some patients had more than one comorbidity.

Healthy controls (n=114) were included from ongoing studies coordinated by the Norwegian Centre for Mental Disorders Research (NORMENT). The controls were scanned with the same MRI scanner and protocol as the NT1 patients. Exclusion criteria for healthy controls were somatic, neurological or psychiatric disease, severe psychiatric family history, metal implants, previous head injury with 30 minutes amnesia or loss of consciousness for 10 minutes, and neuroradiological findings indicating ongoing or previous disease or abnormalities.

### 2.2 Narcolepsy diagnosis

An experienced neurologist and somnologist (S.K.H.) evaluated the patientś clinical and sleep parameters and CSF hypocretin-1 measurements and verified their NT1 diagnosis based on the ICSD-3 criteria. [23] In addition to semi-structured interviews, including a Norwegian translation of the validated Stanford Sleep Questionnaire [24] patients underwent clinical assessments including a neurological examination, human leukocyte antigen (HLA)-typing, routine blood parameter sampling, actigraphy, polysomnography (PSG) and a multiple sleep latency test (MSLT). Spinal taps for the patients were done at their local hospitals prior to or after inclusion. The CSF hypocretin-1 levels were analyzed at the Hormone Laboratory of Oslo University Hospital using a slight modification to the method of Phoenix Pharmaceutical St. Joseph, MO as previously published. [25, 26]

### 2.3 Polysomnography recordings

An actigraphy recording (Philips Actiwatch, Respironics Inc., Murrysville, PA, USA) was performed prior to the polysomnography (PSG) for most patients; 34 patients for 12-14 days (one patient not consistently), 14 patients for 8-11 days (three patients not consistently), two patients wore the actigraphy very inconsistently and 5 patients were without actigraphy. The polysomnography recordings were obtained with the SOMNOmedics system (SOMNOmedics GmbH, Randersacker, Germany) with the electrodes F3-A2, C3-A2, O1-A2, F4-A1, C4-A1 and O2-A1, vertical and horizontal electro-oculography, surface electromyography of the tibialis anterior and submentalis muscles, nasal pressure transducer, nasal thermistor, thoracic respiratory effort, oxygen saturation and electrocardiography. Impedance was kept below 10kΩ (preferably 5Ω). The polysomnography was followed the next day by a 5-nap multiple sleep latency test, where the naps (30 minutes) were separated by 2-hour intervals. The American Academy of Sleep Medicine scoring criteria were used. [23]

### 2.4 MRI acquisition and processing

A General Electric Discovery MR750 3T scanner with a 32-channel head coil were used to obtain the T1-weighted MRI data with the following parameters: Voxel size 1 x 1 x 1 mm; echo time (TE): 3.18 ms; repetition time (TR): 8.16 ms; flip angle: 12°; 188 sagittal slices.

FreeSurfer [27] (v.7.1.0) was used to process the T1-weighted data using the standard recon-all pipeline. FreeSurfer’s Euler number [28] was used for quality control. Datasets with Euler number ± 3 standard deviations from the mean were visually inspected. We obtained three global, 10 bilateral subcortical volume measures and 34 bilateral cortical thickness measures. [29] Bilateral measures were chosen to limit the number of tests in this MRI screening. The bilateral volume measures were computed as the sum of the right and left hemisphere measurement. The bilateral thickness measures were calculated with the formula: Bilateral thickness = (((lh.thickness * lh.surfacearea) + (rh.thickness * rh.surfacearea)) / (lh.surfacearea + rh.surfacearea)).

Previously we have reported on the same 54 patients and 114 controls in a T1-study with segmentation of the hypothalamus [10] and on DTI data [30] for 51 of the patients and fMRI data [31] for 37 of the patients obtained in the same scanning session.

### 2.5 Statistical analysis

We used Permutation Analysis of Linear Models (PALM) [32–34] to test for group differences in volume and thickness. We included age, sex and intracranial volume (ICV) in general linear models for regional volume measures, and age and sex in the models for global volume and thickness measures. We performed 5000 permutations, yielding t-statistics, contrast of parameter estimate (COPE)s, two-tailed p-values and Cohen’s d. Cohen’s d is calculated as COPE / square root (variance of the residuals) in PALM. Due to a sibling pair in the patient group, exchangeability was constrained for these two to control for the lack of independence. We corrected for multiple testing (47 tests - testing group difference in 13 bilateral volume and 34 bilateral thickness measures) with the Benjamini-Hochberg procedure [35] to control the false discovery rate (FDR) at 5%.

Further, for the main models revealing a significant group difference an additional sensitivity analysis was performed, excluding patients with the comorbidities listed above and/or AHI ≥ 5. This additional analysis included 36 patients (27 females, mean age 21.0 ± 11.2) and 114 controls (77 females, mean age 23.2 ± 9.0).

### 2.6 Data availability

Due to ethical approval restrictions, the data are not publicly available as it could compromise the privacy of the participants. NT1 is a rare disorder and Norway has a relatively small population, releasing information about combinations of variables could make some of the participants identifiable.

## 3. Results

### 3.1 Cortical thickness

Table 2 summarizes the uncorrected (not corrected for age and sex) right, left and bilateral mean cortical thickness measures for patients and controls for all the included regions. Permutation testing revealed that patients had thinner cortex bilaterally compared to controls in the temporal poles (*d*=0.68, *p*=0.00080), entorhinal cortex (*d*=0.60, *p*=0.0018), superior temporal gyrus (STG) (*d*=0.60, *p*=0.0020), middle temporal gyrus (*d*=0.50, *p*=0.0088), rostral middle frontal gyrus (*d*=0.49, *p*=0.009), superior frontal gyrus (*d*=0.41, *p*=0.029) and postcentral gyrus (*d*=0.38, *p*=0.043). After FDR correction the bilateral temporal poles, entorhinal cortex and STG remained significant (Figures 1-2).

**Figure 1.**
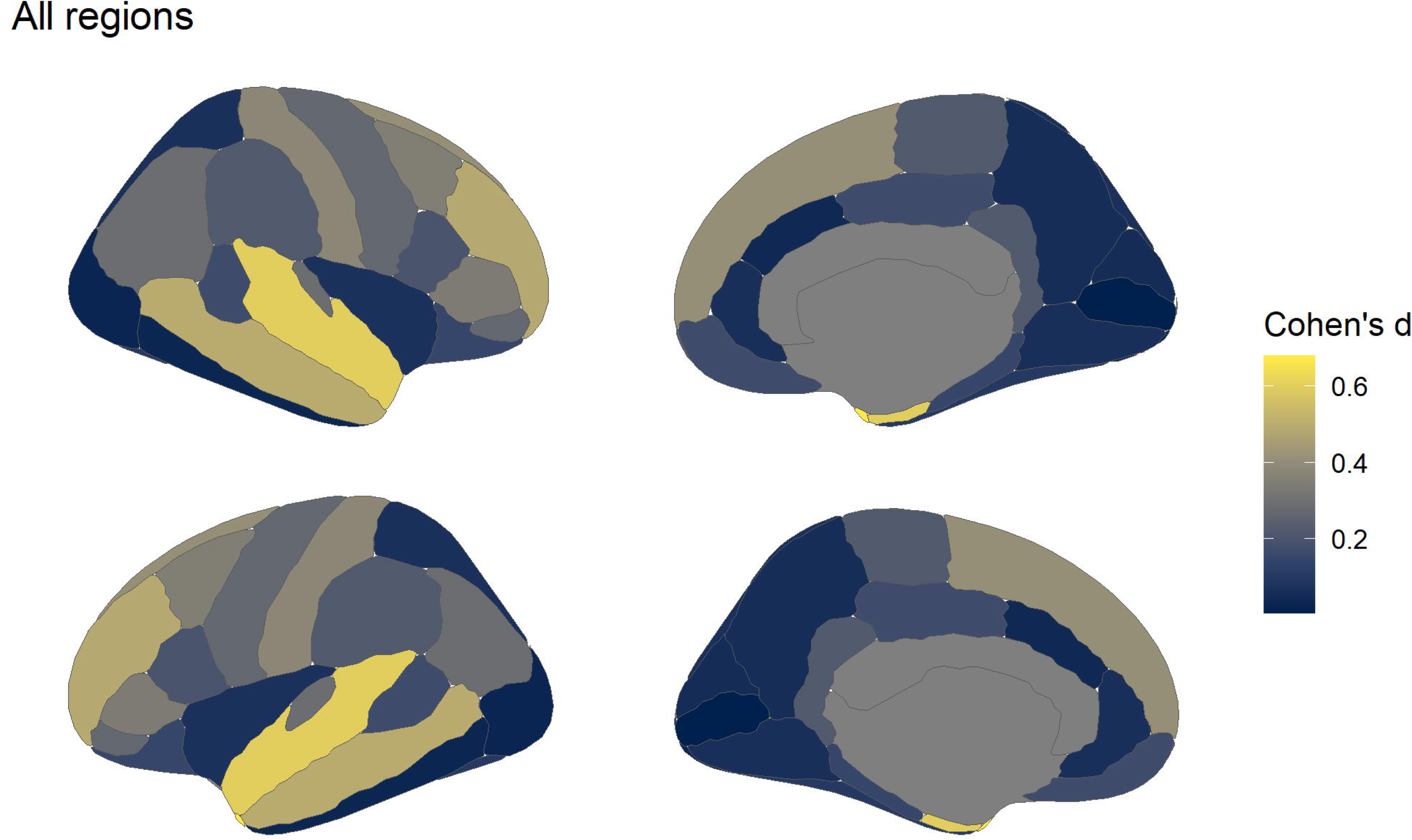
All tested cortical regions. Effect sizes are shown for the thickness measures obtained for 34 bilateral cortical regions. The brain regions are based on the Desikan-Killiany atlas as included in FreeSurfer. [27] Corpus callosum was not a part of this analysis.

**Figure 2.**
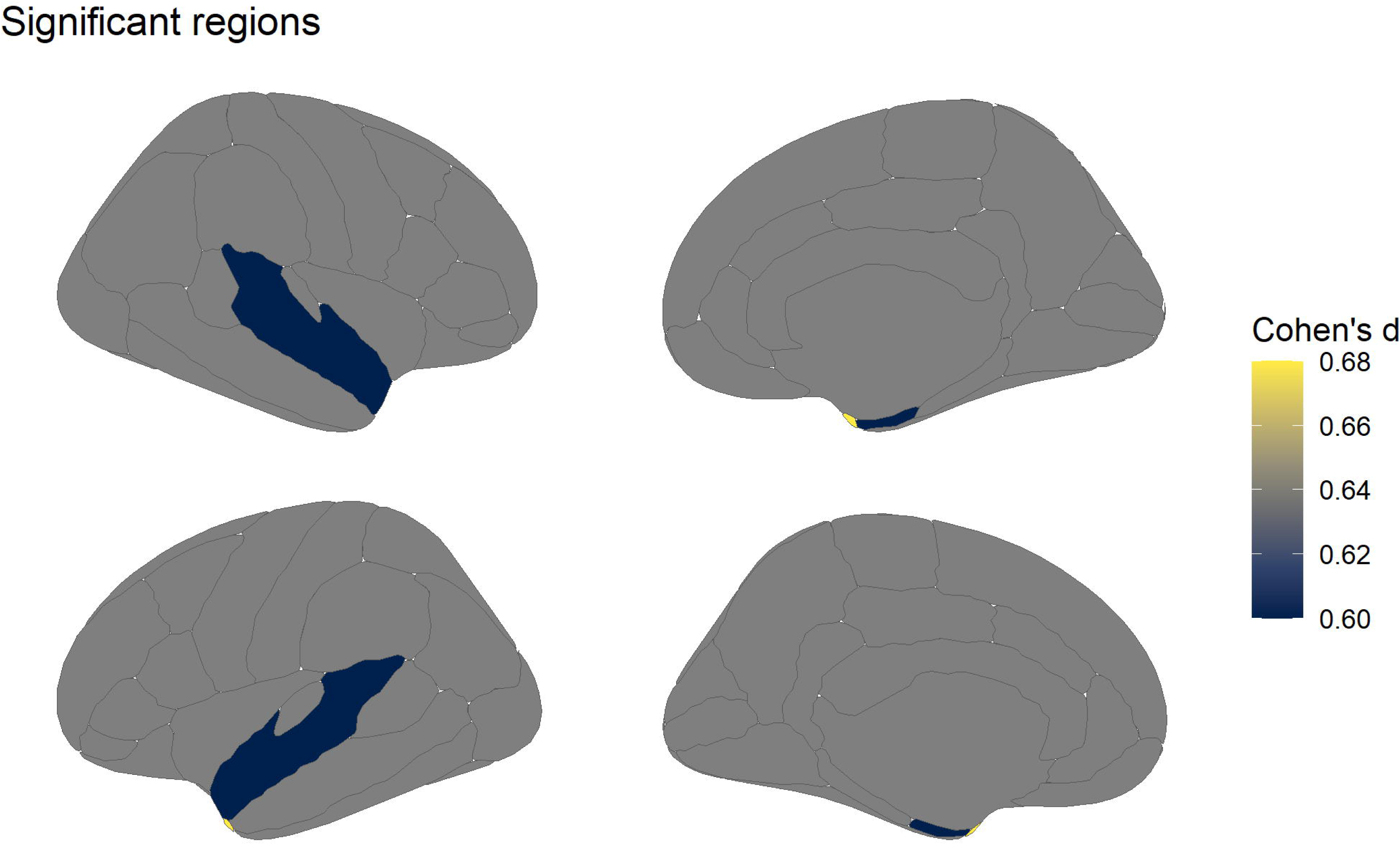
All significant regions. Effect sizes for the three bilateral cortical regions (superior temporal gyrus, entorhinal cortex and temporal poles) where patients had significantly thinner cortex than healthy controls after permutation testing and FDR correction. The brain regions are based on the Desikan-Killiany atlas as included in FreeSurfer. [27]

**Table 2.** Uncorrected mean cortical thickness.

| | NT1 patients (mean ± SD) | | | Controls (mean ± SD) | | | | |
| --- | --- | --- | --- | --- | --- | --- | --- | --- |
|  | Right | Left | Bilateral | Right | Left | Bilateral | p | Cohen's d |
| Temporal poles | 3.25 ± 0.32 | 3.20 ± 0.32 | 3.23 ± 0.28 | 3.38 ± 0.33 | 3.42 ± 0.32 | 3.40 ± 0.29 | 0.00080** | 0.68 |
| Entorhinal | 3.08 ± 0.28 | 3.03 ± 0.28 | 3.04 ± 0.23 | 3.21 ± 0.31 | 3.17 ± 0.27 | 3.18 ± 0.25 | 0.0018** | 0.60 |
| Superior temporal | 2.79 ± 0.15 | 2.84 ± 0.14 | 2.82 ± 0.13 | 2.84 ± 0.14 | 2.90 ± 0.14 | 2.87 ± 0.13 | 0.0020** | 0.60 |
| Middle temporal | 2.77 ± 0.13 | 2.88 ± 0.15 | 2.82 ± 0.12 | 2.80 ± 0.14 | 2.93 ± 0.15 | 2.86 ± 0.13 | 0.0088* | 0.50 |
| Rostral middle frontal | 2.34 ± 0.11 | 2.37 ± 0.13 | 2.36 ± 0.10 | 2.37 ± 0.11 | 2.40 ± 0.12 | 2.39 ± 0.10 | 0.009* | 0.49 |
| Superior frontal | 2.76 ± 0.13 | 2.79 ± 0.13 | 2.78 ± 0.12 | 2.80 ± 0.14 | 2.81 ± 0.15 | 2.81 ± 0.14 | 0.029* | 0.41 |
| Postcentral | 2.13 ± 0.13 | 2.12 ± 0.11 | 2.12 ± 0.11 | 2.16 ± 0.13 | 2.16 ± 0.13 | 2.16 ± 0.12 | 0.043* | 0.38 |
| Banks of the STS | 2.60 ± 0.17 | 2.59 ± 0.16 | 2.60 ± 0.15 | 2.62 ± 0.16 | 2.60 ± 0.15 | 2.61 ± 0.14 | 0.36 | 0.17 |
| Caudal middle frontal | 2.59 ± 0.12 | 2.62 ± 0.17 | 2.61 ± 0.13 | 2.63 ± 0.13 | 2.63 ± 0.14 | 2.63 ± 0.11 | 0.061 | 0.35 |
| Cuneus | 2.07 ± 0.14 | 2.05 ± 0.14 | 2.06 ± 0.13 | 2.06 ± 0.13 | 2.06 ± 0.12 | 2.06 ± 0.12 | 0.76 | 0.055 |
| Insula | 3.07 ± 0.17 | 3.09 ± 0.17 | 3.08 ± 0.16 | 3.05 ± 0.14 | 3.07 ± 0.14 | 3.06 ± 0.13 | 0.72 | 0.070 |
| Inferior temporal | 2.71 ± 0.15 | 2.77 ± 0.18 | 2.74 ± 0.15 | 2.69 ± 0.13 | 2.78 ± 0.14 | 2.74 ± 0.12 | 0.87 | 0.031 |
| Caudal anterior cingulate | 2.46 ± 0.19 | 2.52 ± 0.19 | 2.49 ± 0.17 | 2.46 ± 0.17 | 2.49 ± 0.18 | 2.48 ± 0.15 | 0.84 | 0.039 |
| Posterior cingulate | 2.52 ± 0.13 | 2.48 ± 0.16 | 2.50 ± 0.13 | 2.53 ± 0.13 | 2.48 ± 0.14 | 2.51 ± 0.12 | 0.38 | 0.17 |
| Fusiform | 2.79 ± 0.15 | 2.80 ± 0.12 | 2.80 ± 0.13 | 2.79 ± 0.12 | 2.81 ± 0.11 | 2.80 ± 0.11 | 0.60 | 0.099 |
| Inferior parietal | 2.45 ± 0.14 | 2.44 ± 0.12 | 2.45 ± 0.11 | 2.47 ± 0.13 | 2.47 ± 0.13 | 2.47 ± 0.12 | 0.12 | 0.29 |
| Isthmus cingulate | 2.47 ± 0.18 | 2.43 ± 0.16 | 2.45 ± 0.16 | 2.43 ± 0.16 | 2.41 ± 0.16 | 2.42 ± 0.14 | 0.27 | 0.22 |
| Lateral occipital | 2.20 ± 0.10 | 2.16 ± 0.10 | 2.18 ± 0.09 | 2.20 ± 0.12 | 2.16 ± 0.11 | 2.18 ± 0.11 | 0.87 | 0.028 |
| Lateral orbitofrontal | 2.55 ± 0.15 | 2.57 ± 0.15 | 2.56 ± 0.14 | 2.57 ± 0.12 | 2.57 ± 0.14 | 2.57 ± 0.11 | 0.41 | 0.15 |
| Lingual | 2.24 ± 0.15 | 2.23 ± 0.12 | 2.24 ± 0.13 | 2.21 ± 0.13 | 2.23 ± 0.12 | 2.22 ± 0.12 | 0.72 | 0.066 |
| Medial orbitofrontal | 2.45 ± 0.13 | 2.46 ± 0.15 | 2.46 ± 0.13 | 2.44 ± 0.11 | 2.44 ± 0.12 | 2.44 ± 0.10 | 0.38 | 0.17 |
| Parahippocampal | 2.80 ± 0.22 | 2.86 ± 0.18 | 2.83 ± 0.17 | 2.84 ± 0.19 | 2.87 ± 0.26 | 2.85 ± 0.20 | 0.43 | 0.15 |
| Paracentral | 2.57 ± 0.14 | 2.56 ± 0.14 | 2.56 ± 0.13 | 2.60 ± 0.13 | 2.56 ± 0.12 | 2.58 ± 0.12 | 0.24 | 0.22 |
| Pars opercularis | 2.65 ± 0.15 | 2.69 ± 0.13 | 2.67 ± 0.12 | 2.66 ± 0.14 | 2.69 ± 0.13 | 2.68 ± 0.12 | 0.29 | 0.20 |
| Pars orbitalis | 2.68 ± 0.15 | 2.76 ± 0.16 | 2.72 ± 0.14 | 2.72 ± 0.17 | 2.76 ± 0.20 | 2.74 ± 0.16 | 0.14 | 0.27 |
| Pars triangularis | 2.51 ± 0.15 | 2.53 ± 0.16 | 2.52 ± 0.13 | 2.55 ± 0.14 | 2.55 ± 0.14 | 2.55 ± 0.12 | 0.075 | 0.34 |
| Pericalcarine | 1.76 ± 0.15 | 1.89 ± 0.16 | 1.82 ± 0.13 | 1.75 ± 0.18 | 1.89 ± 0.14 | 1.82 ± 0.15 | 0.98 | 0.0043 |
| Precentral | 2.55 ± 0.13 | 2.59 ± 0.13 | 2.57 ± 0.12 | 2.59 ± 0.12 | 2.59 ± 0.13 | 2.59 ± 0.11 | 0.15 | 0.27 |
| Precuneus | 2.57 ± 0.12 | 2.49 ± 0.11 | 2.53 ± 0.11 | 2.56 ± 0.11 | 2.50 ± 0.11 | 2.53 ± 0.11 | 0.76 | 0.058 |
| Rostral anterior cingulate | 2.92 ± 0.23 | 2.87 ± 0.20 | 2.89 ± 0.18 | 2.89 ± 0.20 | 2.85 ± 0.16 | 2.87 ± 0.15 | 0.74 | 0.064 |
| Superior parietal | 2.30 ± 0.10 | 2.28 ± 0.09 | 2.29 ± 0.09 | 2.30 ± 0.11 | 2.28 ± 0.11 | 2.29 ± 0.10 | 0.73 | 0.068 |
| Supramarginal | 2.58 ± 0.14 | 2.50 ± 0.17 | 2.53 ± 0.13 | 2.59 ± 0.14 | 2.53 ± 0.16 | 2.55 ± 0.13 | 0.25 | 0.22 |
| Frontal poles | 2.63 ± 0.24 | 2.71 ± 0.24 | 2.66 ± 0.19 | 2.67 ± 0.31 | 2.72 ± 0.25 | 2.69 ± 0.24 | 0.20 | 0.24 |
| Transverse temporal | 2.57 ± 0.21 | 2.54 ± 0.23 | 2.55 ± 0.20 | 2.61 ± 0.19 | 2.56 ± 0.20 | 2.58 ± 0.17 | 0.12 | 0.29 |

Thickness(mm) is not corrected for age or sex. Two-tailed p-values and the Cohen's d are from the analysis of the bilateral values with models including correction for age and sex. Bilateral thickness measures were calculated with the formula: Bilateral thickness = $((lh.thickness * lh.surfacearea) + (rh.thickness * rh.surfacearea)) / (lh.surfacearea + rh.surfacearea)$ . NT1 patients: narcolepsy type 1 patients, SD: standard deviation, STS: Superior temporal sulcus. \*Significant after permutation testing, \*\* significant after permutation testing and correction for multiple testing with the Benjamini-Hochberg-procedure.

### 3.2 Subcortical and global brain volumes

Table 3 summarizes the uncorrected (not corrected for age, sex or ICV) mean for right, left and bilateral cortical and sub-cortical volumes for patients and controls for all the tested regions. Compared to controls, patients showed smaller global cortical volume (*d*=0.43, *p*=0.03) and smaller total gray matter (GM) volume (*d*=0.37, *p*=0.059). However, after FDR correction these differences were not significant.

**Table 3.**
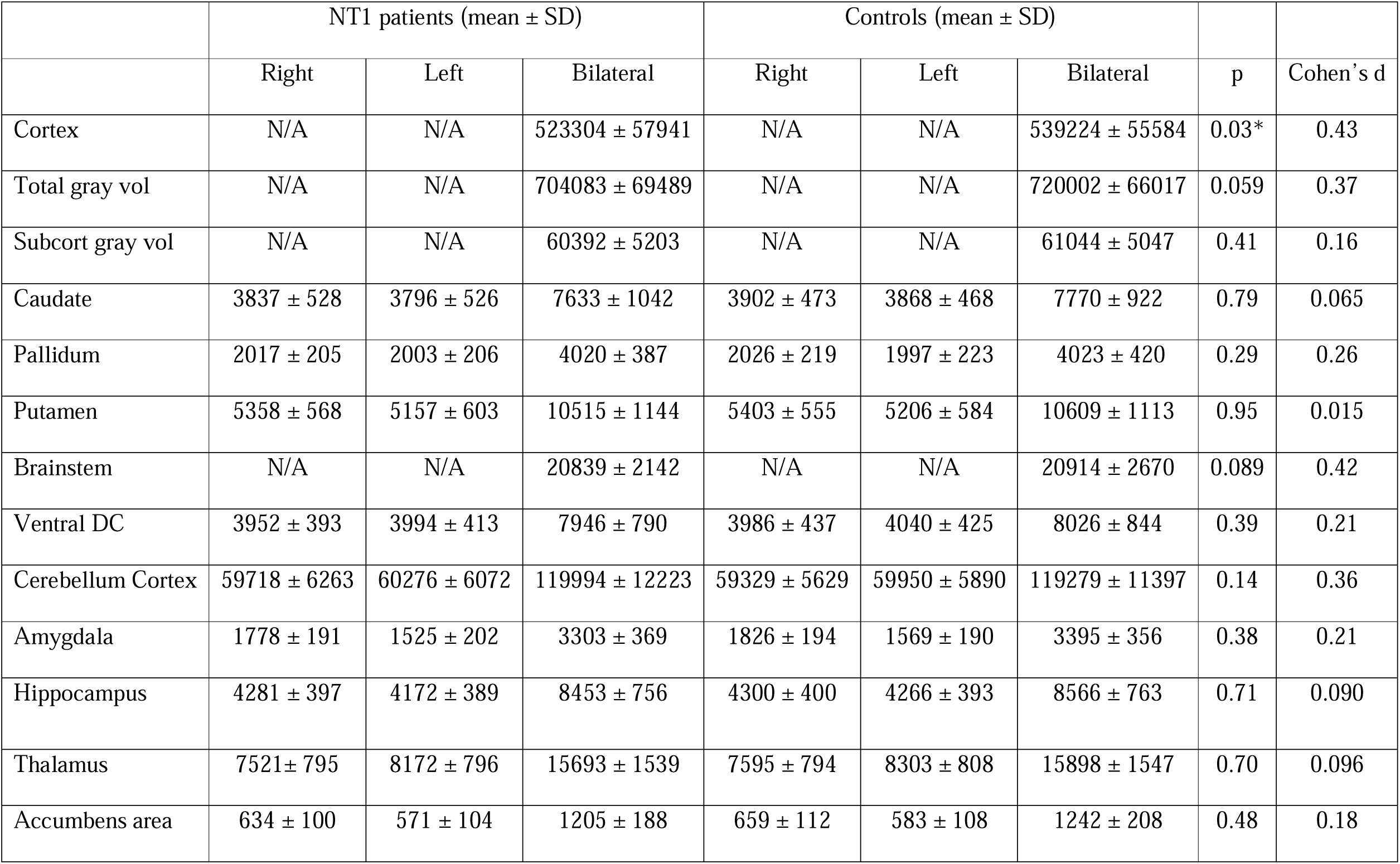
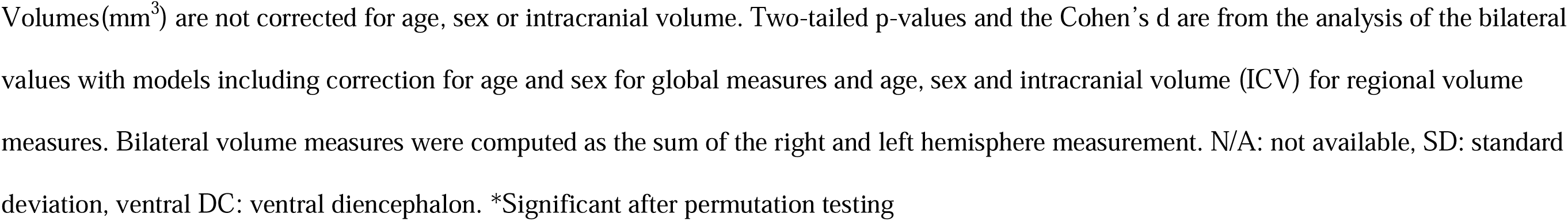
Uncorrected mean volume.

### 3.3 Sensitivity analysis

Sensitivity analyses were performed for the bilateral temporal poles, entorhinal cortex and STG thickness measures, excluding patients with the comorbidities listed above and AHI ≥5. Permutation testing revealed similar results as in the main analysis with thinner cortex bilaterally for patients compared to controls for the temporal poles (*d*=0.61, *p*=0.0088), entorhinal cortex (*d*=0.58, *p*=0.013) and STG (*d*=0.48, *p*=0.032).

## 4. Discussion

Our main finding was significantly thinner cortex for NT1 patients compared to controls bilaterally in the STG, temporal poles and entorhinal cortex, all located in the temporal lobe. Several previous studies [11, 12, 14, 18, 36] have implicated changes in the STG for narcolepsy patients, but our study is the first to report thinner cortex in the temporal poles and entorhinal cortex.

The STG is critical for speech perception and injuries to this region lead to extensive deficits in the comprehension of speech. [37] Wernicke’s aphasia after damage to the STG is characterized by fluent speech, but severely impaired comprehension. [38] The STG is considered to encode the more complex features of sound than the earlier regions of the auditory hierarchy and is therefore considered to be a part of the higher-order associative auditory cortex. [37] The STG has been proposed as a key area for structural pathology and a possible biomarker for schizophrenia (SZ) patients. [39] Electrical stimulation of the STG can cause hallucinations and it is thought to play a role in auditory hallucinations and thought disorder in SZ. [39, 40] Smaller STG volumes in SZ patients compared to controls has been reported in several studies [39] and a longitudinal study of patients with first-episode SZ suggested a progressive volume reduction of the left posterior STG 1.5 years after the first hospitalization. [41]

In our study 85 % of the NT1 patients have experienced hypnagogic (at sleep onset) or hypnopompic (on awakening) hallucinations. Likewise, other studies have indicated that up to 80% of NT1 patients experience hypnagogic/hypnopompic hallucinations, but some also report daytime hallucinations not connected to sleep. [42] The hallucinations in NT1 patients are presumed to represent intrusion of REM-sleep into wakefulness. [42] Most often patients are aware that the hallucinations are not real, however it has been reported that differentiating hallucinations from reality can be a struggle for otherwise mentally healthy narcolepsy patients. [42, 43] Unlike SZ patients who often have verbal-auditory hallucinations, multisensory (visual, auditory, olfactory and tactile) hallucinations dominate for NT1 patients. [42, 43] However, NT1 patients can have more non-verbal auditory hallucinations (footsteps, animal sounds etc.) than SZ patients and there has been cases where the hallucinations in NT1 patients have led to a misdiagnosis of SZ. [1, 42] Only eight of our NT1 patients had not experienced hypnagogic/hypnopompic hallucinations, and the associations between hallucinations and cortical thickness among NT1 patients should be explored in a larger multi-center study to gain enough patients without hallucinations.

The observed group differences might also be attributed to secondary effects due to abnormal sleep-wake patterns. Compared to day-workers reduced STG volume has been reported among shift workers who also reported more sleep disturbances and depressive symptoms. [44] Further, thinner STG has also been reported in patients with obstructive sleep apnea (OSA). [45] In our study, six NT1 patients had AHI ≥ 5, however these patients were excluded from the sensitivity analysis which largely supported the results from the main analysis.

Interestingly we also found significantly thinner cortex of the temporal poles for the patients compared to controls in the present study, which has not been reported previously. The temporal poles are thought to play a role in memory and a new class of face memory cells was reported in a recent study [46] from recordings of the temporal pole in rhesus monkeys. These cells do not respond to faces in general, but to faces that are personally familiar. [46] The temporal pole has also been associated with visual processing of complex objects, naming and word object labeling and autobiographic memory. [47] It has also been revealed that the temporal pole has a role in language and semantic processing, with activation in PET studies during linguistic tasks as for example recall of word lists and story recall. [47] Furthermore, the temporal poles have been associated with socio-emotional processing. [47] An example of this can be seen with a variant of frontotemporal lobar degeneration (FTLD) which gives right temporal pole atrophy and have symptoms like social awkwardness, difficulty in identifying people, loss of insight, loss of empathy, aggressive behavior, irritability, poor emotional regulation and personality changes. [47] In patients with traumatic brain injury impairment of a social problem-solving task was associated with cortical thickness of the temporal pole. [47] Although the overall clinical picture is clearly different in NT1 compared to for example FTLD, some degree of social problems are frequently reported in NT1, and a study [48] including 53 children with NT1 found that 45% had a relevant social impairment, even if they all received treatment and 70% were medicated.

A large meta-analysis [49] of 4474 SZ patients and 5098 healthy controls reported a stronger negative correlation between age and cortical thickness in the bilateral temporal poles for the patients than the controls. Further, a longitudinal study [50] of first-episode SZ patients showed smaller GM volume and progressive GM volume decrease in the temporal pole after 1.5 years. Future longitudinal studies should explore if a progressive decrease of the temporal poles and the STG can be found in NT1 patients as well.

We also found significantly thinner entorhinal cortex in NT1 patients compared to controls, which has not previously been reported. The entorhinal cortex is important for episodic memories, which can be defined as the ability to recall what happened in a spatiotemporal context, [51–53] and input from the entorhinal cortex to the hippocampal formation is a critical part of being able to form these types of memories. [53] The entorhinal cortex is also important for spatial navigation and the forming of universal maps, and in the medial entorhinal cortex spatially modulated cells can be found like the grid cells. [51–53]

The entorhinal cortex has also been implicated in an MRI study of OSA patients, investigating structural MRI changes in relation to cognitive deficits before and after treatment (3 months with continuous positive airway pressure (CPAP)). [54] Before treatment OSA patients exhibited smaller GM volumes compared to controls in several regions including the entorhinal cortex. Entorhinal volume increase after treatment was correlated with improvements in tests for short-term memory and executive functions. [54] The authors speculated that structural and cognitive deficits in OSA patients might be secondary to the hypoxemia, disturbances in autonomic activity or the sleep fragmentation. [54] NT1 patients have problems with maintaining sleep and wakefulness, [1] which leads to sleep fragmentation and could partly explain our findings and provide a possible link to MRI findings in OSA. As mentioned, six NT1 patients in our study have AHI ≥ 5, but as they were excluded from the sensitivity analysis, which largely supported the results from the main analysis, it is unlikely that this can explain the group difference.

A recent study compared performance on a visual perceptual learning task before and after naps between 15 NT1 patients, 14 Narcolepsy type 2 (NT2) patients (hypocretin-1 levels are per definition normal in the CSF of NT2 patients [23]) and 7 idiopathic hypersomnia patients. [55] NT1 patients often have naps starting in REM-sleep and then continuing to non-REM-sleep (NREM), and the study found that sleep spindles only were associated with memory consolidation when the naps started with NREM-sleep followed by REM-sleep. [55] However, studies have indicated that possible memory deficits in NT1 patients seem to be limited to specific memory functions and there have been some indications of alterations of procedural memory with less effective memory consolidation as well as impaired working memory. [56]

Previous structural MRI studies of narcolepsy patients [11–22] have revealed conflicting results involving various brain regions. While most of the studies have been brain-wide [11–22]some studies have looked at regions of interest in narcolepsy patients like amygdala [57–59], hippocampus [57, 58, 60] and nucleus accumbens [59]. To the best of our knowledge our study represents the first brain-wide T1-study of a post-H1N1(largely Pandemrix®-vaccinated) patient sample. Previous studies do not contain information in regards to H1N1-vaccination status. However, as far as we can tell previous studies are either of sporadic narcolepsy samples before the H1N1-vaccination had occurred [11–18, 20], or have patients samples [19, 21, 22] from countries that did not use Pandemrix® like South-Korea [61] and Italy [8].

Although the results from the previous studies are conflicting a review from 2019 [62] including 13 structural MRI studies [11–20, 57–59, 63] on narcolepsy showed that replicated findings (defined as being reported in two or more articles) were reductions of superior frontal gyri, superior and inferior temporal gyri, middle occipital gyri, hypothalamus, amygdala, insula, hippocampus, cingulate cortex, thalamus and nucleus accumbens. However, the authors stated that the replicated findings are controversial due to several issues, including small and heterogeneous patient samples with large variation in age and disease duration, which means that effect of aging, illness and possible compensatory mechanisms are difficult to distinguish.

A meta-analysis [36] of eight voxel-based morphometry studies [11–18] of narcolepsy patients where the studies either assessed GM concentration [11, 14, 18] or GM volume [12, 13, 15–17] reported significant GM reductions in patients bilaterally in the hypothalamus, nucleus accumbens, anterior cingulate cortex, thalamus, left mid orbital and rectal gyri, right inferior frontal and superior temporal gyri. However, the authors of the meta-analysis [36] pointed out several limitations as the included studies applied different inclusion and exclusion criteria, heterogenous patient samples and methodologies. In addition, three additional meta-analysis [64–66] have been performed on the same data as Weng *et al*. [36] with conflicting results. The first meta-analysis [66] being a re-analysis due to a software correction that had occurred after Weng *et al*. [36] had published, concluded that they could not replicate the findings from Weng *et al.* [36] and that therefore there was no evidence of spatially consistent localized loss of grey matter in narcolepsy. Another meta-analysis [64] then used another technique including the same data as Weng *et al.* [36] and had the most robust findings of grey matter atrophy in bilateral superior frontal gyrus and hypothalamus, striatum and thalamus. However, the most recent meta-analysis [65] stated that Zhong *et al*. [64] had not controlled the type 1 error rate sufficiently, and that some studies violated the inclusion criteria for the meta-analysis. Tench *et al*. [65] performed another re-analysis of the data and found again no evidence for spatially consistent effects. Although Tench *et al*. [65] acknowledged limitations with the meta-analysis when there are so few studies included and therefore only having statistical power to detect strong effects, and that additionally the risk of false positive results increases due to small sample sizes in some of the studies and that half of the studies reported uncorrected p-values. [65]

Our study is the first study to report thinner cortex in the temporal poles and entorhinal cortex in patients compared to controls, but as previously mentioned several studies [11, 12, 14, 18, 36] have implicated the superior temporal regions in NT1. The difference in findings could possibly be related to our NT1 patient sample being post-H1N1 (largely Pandemrix®-vaccinated). It has since the 2009 H1N1-vaccinations been an ongoing discussion if the previously known sporadic NT1 and NT1 after H1N1-vaccination are exactly the same disease entity/phenotype, although most studies point in to the “one entity/phenotype” direction [7, 67–69] some possible differences has been reported like for example a sudden onset of symptoms [6, 7, 69–74], more disturbed sleep characteristics [72, 74], and case reports of severe psychiatric symptoms in narcolepsy linked to the Pandemrix®-vaccination [74]. Furthermore, in a recent T1-study [10] by our group of the same 54 post-H1N1 patients and 114 controls as included in the current study we explored volume differences of the hypothalamus with a segmentation tool included in Freesurfer and revealed significantly larger hypothalamic volume, particularly in the tubular-inferior subregions in patients compared to controls. The reported increased volume of hypothalamus could possibly reflect gliosis, neuroinflammation or changes in cell types as previously mentioned and it cannot be ruled out that this hypothalamic volume increase that have not previously been found in NT1 patients is specific to post-H1N1(largely Pandemrix-vaccinated) NT1. [10]

In another previous study by our group using diffusion tensor imaging [30] we found widespread white matter changes affecting 34.2% of the white matter skeleton in post-H1N1 (largely Pandemrix®-vaccinated) NT1 patients (involving 51 of the patients included in the current study), which were far more widespread than previously reported in sporadic narcolepsy [30]. Although our DTI-study had almost three times more patients than the largest study of sporadic narcolepsy [75] and this increase in power could also be part of the explanation for the difference in results. Similarly, the current study has almost twice the sample size (n = 54) as the largest previous study (n=29 narcolepsy patients) [11] investigating brain volume across a large number of brain regions, which reduces the uncertainty of the effect sizes and the risk of both false negatives and false positives

Another possible explanation for the difference in findings could be related to our patient sample being homogenous and well-characterized with verified hypocretin-deficiency in 51/54 patients (three NT1 patients had not yet had their hypocretin values measured, but had cataplexy, positive PSG/MSLTs, and were HLA-DQB1*0602-positive) and a disease duration of approximately 6 years. The largest previous study [11] did not report information about disease duration, medication and hypocretin-deficiency status and it was unclear if both NT1 patients and NT2 patients were included. Although the information is not available for every article, many of the previous studies have large variation in disease duration, from 1-40 years [17], 3 -55 years [16], 2–50 years [13] and 6–-59 years [18]. It could also be related to most of the previous studies [11–14, 16–19, 59] using 1.5-T scanners except for four studies. [15, 20, 21, 58].

Comorbidities are frequently observed among patients with narcolepsy, [2] and this may have confounded the initial case-control comparisons. However, sensitivity analysis largely supported the main findings, suggesting that the group differences cannot be simply explained by known comorbid conditions.

In conclusion, compared to healthy controls, hypocretin-deficient post-H1N1(largely Pandemrix®-vaccinated) NT1 patients have thinner MRI-based cerebral cortex in several temporal brain regions, including bilateral STG, entorhinal cortex and temporal poles. Entorhinal cortex and temporal poles are involved in memory functions and STG play a role in speech and hallucinations. Although speculative, these findings may reflect secondary effects of a primary loss/dysfunction of the hypocretin-producing neurons. Alternatively, the findings might be associated with the characteristic abnormal sleep-wake pattern in NT1 patients, or they could be specific to post-H1N1 NT1(largely Pandemrix®-vaccinated) patients. Future longitudinal studies are needed to assess the clinical relevance and dynamics of the observed brain structural abnormalities.

## Data Availability

Due to ethical restrictions the data are not publicly available as it could compromise the privacy of the participants, as NT1 is a rare disorder and Norway has a relatively small population.

## 5. Acknowledgments

A preprint of the paper has been posted on medRxiv. A special thank you goes to the participating patients. We also want to thank Janita Vevelstad (nurse at the Norwegian Centre of Expertise for Neurodevelopmental Disorders and Hypersomnias (NevSom), Oslo University Hospital) and Rannveig Viste (nurse, molecular biologist and PhD-student at the Norwegian Centre of Expertise for Neurodevelopmental Disorders and Hypersomnias (NevSom), Oslo University Hospital) for being a part of the data collection and sleep scoring of polysomnography/multiple sleep latency tests. Furthermore, we would like to express our appreciation for Ranveig Østrem’s (bioengineer at the Hormone Laboratory at Oslo University Hospital) work with the HLA-typing. **Funding:** S. K. H was partially funded by research support from the Norwegian Ministry of Health and Care Services. H.T.J was during data collection fully funded by the Norwegian Ministry of Health and Care Services and during data analyses and manuscript writing by the Helse SØ grant (2019032). D. A was supported by the South-Eastern Norway Regional Health Authorities (2019107, 2020086). The European Union’s Horizon 2020 Research and Innovation program (ERC StG, Grant # 802998), the Research Council of Norway (223273, 249795, 300767) and the South-Eastern Norway Regional Health Authority (2019101) was also part of funding this study. The funding sources had no involvement with the study design, data collection, analysis or interpretation of the data, the writing of the article or in the decision to submit the article for publication.

